# SARS-CoV-2 transmission dynamics in Mozambique and Zimbabwe during the First Three Years of the Pandemic

**DOI:** 10.1101/2024.05.20.24307570

**Authors:** Roselyn F Kaondera-Shava, Marta Galanti, Matteo Perini, Jiyeon Suh, Shannon M Farley, Sergio Chicumbe, Ilesh Jani, Annette Cassy, Ivalda Macicame, Naisa Manafe, Wafaa El-Sadr, Jeffrey Shaman

## Abstract

The 2019 emergence of severe acute respiratory syndrome coronavirus 2 (SARS-CoV-2) and its rapid spread created a public health emergency of international concern. However, the impact of the pandemic in Sub-Saharan Africa, as documented in cases, hospitalizations and deaths, appears far lower than in the Americas, Europe, and Asia. Characterization of the transmission dynamics is critical for understanding how SARS-CoV-2 spread and the true scale of the pandemic. Here, to better understand SARS-CoV-2 transmission dynamics in two southern African countries, Mozambique and Zimbabwe, we developed a dynamic model-Bayesian inference system to estimate key epidemiologic parameters, namely the transmission and ascertainment rates. Total infection burdens (reported and unreported) during the first three years of the pandemic were reconstructed using a model-inference approach. Transmission rates rose with each successive wave, which aligns with observations in other continents. Ascertainment rates were found to be low matching prior assumption and consistent with other African countries. Overall, the estimated disease burden was higher than the documented cases, indicating need for improved reporting and surveillance. These findings aid understanding of COVID-19 disease and respiratory virus transmission dynamics in two African countries little investigated to date, and can help guide future public health planning and control strategies.

## 1 Introduction

Following its emergence in China in late December 2019, severe acute respiratory syndrome coronavirus 2 (SARS-CoV-2), the virus that causes coronavirus disease 2019 (COVID-19) spread globally and produced an unprecedented public health crisis [44, 49]. The World Health Organization (WHO) declared the COVID-19 outbreak a pandemic on March 11*^th^*, 2020 [8]. In less than three years, in various parts of the world, including two southern African countries, i.e. Mozambique and Zimbabwe, SARS-CoV-2 triggered multiple waves of new infections, often driven by new variants of concern (VOC) [30].

The COVID-19 pandemic put enormous pressure on public health security; to mitigate COVID-19 spread, most African countries imposed travel restrictions or banned international travel and implemented curfews, lockdowns and other social distancing interventions beginning in March and April 2020 [31]. The initial cases of COVID-19 in Mozambique and Zimbabwe were reported on March 22*^nd^* and March 20*^th^* 2020 [20, 35], respectively, and the pandemic subsequently spread to all provinces in both countries. By December 2022, three waves of infection, i.e., Alpha, Delta, and Omicron (BA.1), had occurred.

Characterization of transmission dynamics is critical for understanding how SARS-CoV-2 spread and the full extent of its impact on local populations. Statistical and modeling studies have provided important insights into disease transmission dynamics by estimating critical epidemiological parameters (such as the reproductive number, ascertainment rates, and total infections) in various studies. For example, the time-varying reproductive number characterizes virus transmissibility at a particular point in time; this quantity has been estimated using stochastic models coupled with Bayesian inference for France and Ireland [6], Switzerland [32], and Scandinavian countries [19]. Odurro *et al.* [28] used a time varying reproduction estimation approach for sub-Saharan Africa and estimated the time-varying reproductive number while also accounting for depletion of the susceptible population. In addition, a model-inference approach was used to estimate the background population characteristics such as population susceptibility for South Africa [46], the United States at county resolution [34], and India [45].

Many SARS-CoV-2 infections are unreported; hence, confirmed case counts do not fully reflect epidemic dynamics. Understanding the level of ascertainment and the impact of undocumented cases on transmission is crucial to plan and implement control strategies. Models coupled with Bayesian inference methods have been used to investigate the role of undocumented infections in China [22], as well as for various regions (countries and US states) [7], 54 African countries [18], and 3 North African countries: Algeria, Egypt, and Morocco [11]. Russel *et al.* [36] fitted a Bayesian Gaussian process model to estimate under-ascertainment in 210 countries and territories.

In spite of this vast body of literature, characterization of SARS-CoV-2 transmission dynamics (or any respiratory virus) in the Southern African region remains mostly lacking, except for South Africa [46, 40, 48, 25]. An autoregressive integrated moving average (ARIMA) model was utilized to forecast the trend of the disease in four African countries which reported the most cases: South-Africa, Egypt, Nigeria and Ghana [23] and in Southern Africa [38]. Shoko *et al.* [39] used a support vector regression for short-term forecasting of the disease in Zimbabwe. However, the reported COVID-19 impact (cases, hospitalizations, and deaths) in Africa has likely underestimated the actual extent of infection and thus the transmission dynamics. As a consequence, investigation of key epidemiological characteristics over time and by VOC wave is needed.

Here we utilize a model-inference framework to study COVID-19 disease dynamics in Mozambique and Zimbabwe during the first three years of the pandemic, for the three VOCs Alpha, Delta and Omicron (BA.1). The dynamic model was coupled with a Bayesian inference method and province-specific case data. We accounted for undocumented cases to estimate key epidemiologic parameters, namely the transmission rate and the ascertainment rate at the end of each variant outbreak. The findings help advance understanding of the dynamics of SARS-CoV-2 and give information on the transmission dynamics of respiratory viruses in two African nations that have received little attention to date.

## 2 Methodology

### 2.1 Epidemic transmission model

To describe the prevalent characteristics of COVID-19 in each province, *i*, for Mozambique and Zimbabwe, we used a Susceptible-Exposed-Infectious-Recovered (SEIR) compartmental model, similar to ones used in recent studies on COVID-19 [42, 41, 22, 16]. The infected compartment was split into two compartments; infected reported (*I^r^*), individuals who have tested positive, and infected unreported (*I^u^*), individuals who are infected with the virus but have not been tested and thus not reported. The susceptible population, *S_i_*, is not yet infected, the exposed population, *E_i_*, has been infected with the virus but do not yegt transmit the virus. There is no reinfection of the recovered population, *R_i_*, and *N_i_* is the total population.

The model (see Figure 1) was coupled with data assimilation and case data in order to estimate the transmission rate, *β_i_*and the ascertainment rate, *α_i_*, at the end of each variant wave outbreak. The other parameters, i.e., average latency period, *Z*, average duration of infection, *D* and reduction of infection rate for unreported infected individuals, *µ* were kept constant (see Table S5). We also maintained a constant human population for the study period with no births or deaths.

**Figure 1:**
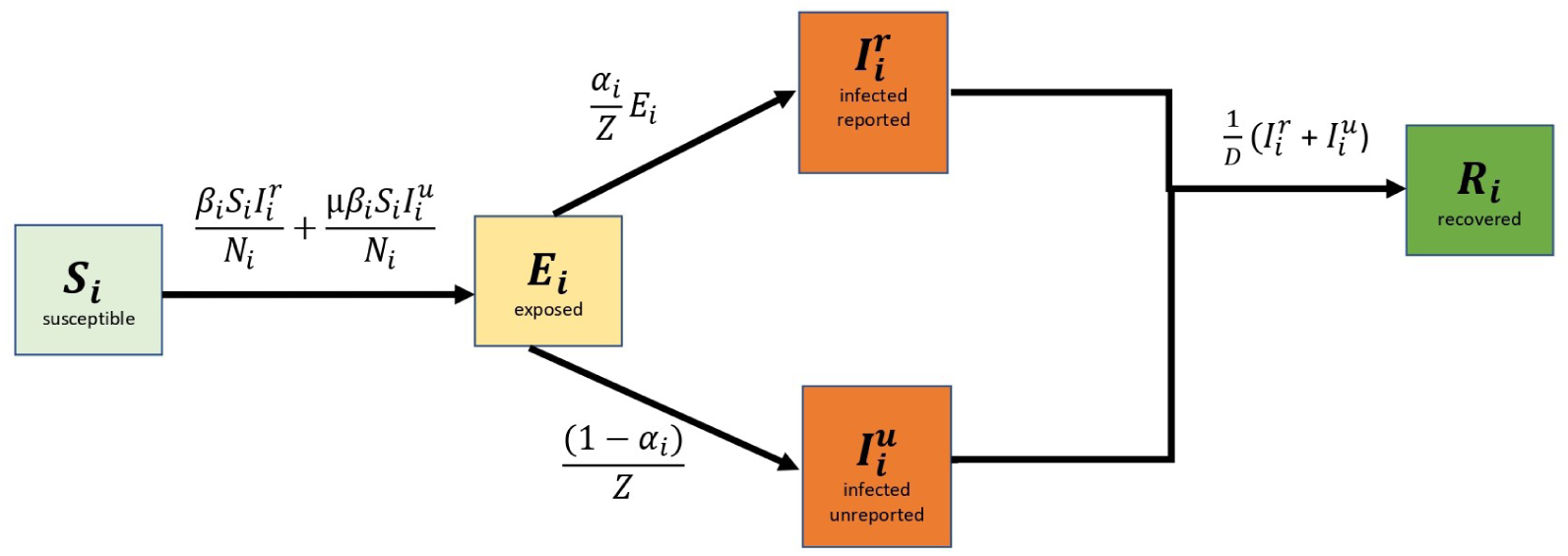
Schematic presentation of simulated disease dynamics. The solid arrows demonstrate the movement from one disease state to another.

#### 2.1.1 Model assumptions

The model is based on the following assumptions:

i. there is no human movement between the provinces
ii. individuals who are infected (reported and unreported) can transmit the virus to those susceptible,
iii. there is no re-infection during each variant wave period.

#### 2.1.2 Disease dynamics

The system of ordinary differential equations was adapted from Li *et al.* [22]. Figure 1 presents a schematic of model transmission dynamics, and a description of the model variables and parameters is shown in Table S3.

Model disease dynamics are described by a system of ordinary differential equations, Equation (S1)–Equation (S4) which are subject to the non-negative initial conditions and satisfy the conservation of the population, i.e., *N_i_* = *S_i_* + *E_i_* + *I_i_^r^* + *I_i_^u^* + *R_i_* for each province *i*.

The reproductive number at a particular time, *Rt_i_* = *α_i_β_i_D*+(1*−α_i_*)*µβ_i_D* was obtained through the next generation approach (see Section S1.3 and Section S1.4 for detailed explanation).

In our analysis, the SEIR model was integrated stochastically forward in time. A Latin hypercube sampling (LHS) technique was used to select a random set of initial variables and parameter combinations from prior ranges, which are assumed to be uniformly distributed. The initial priors for the variables and parameters are given in Table S5. A 4th-order Runge Kutta (RK4) scheme was used for stochastic integration; in particular, to introduce stochasticity to the right hand side of the system, a random sample from a Poisson distribution, was used for each step of the RK4 scheme to determine each term as shown in Equation (S6)–Equation (S9).

For both countries all provinces were run simultaneously as a metapopulation model without representation of human movement between provinces, due to the absence of robust mobility data. Attempted simulations using a gravity model to estimate inter-provincial movement did not yield credible results (described in Section S1.9). As a consequence, we assumed disease transmission dynamics to be principally determined by local within-province processes.

#### 2.1.3 Piecewise Simulation and Inference

The metapopulation model form was coupled with the ensemble adjustment Kalman filter (EAKF), a Bayesian sequential ensemble filtering method, and province-specific COVID-19 case data. We assume no re-infection during each wave; piecewise simulation and parameter estimation were performed separately for each variant wave: Alpha, Delta and Omicron (BA.1). The data periods for this piecewise inference are shown in Table S4. Thus, for each province *i*, we produced parameter estimates for the transmission rate, *β_i_* and ascertainment rate, *α_i_*, for each variant wave period. The procedure for parameter estimation using the EAKF is summarized in Algorithm S1.

The model is reinitialized for each wave and does not explicitly represent re-infections. In Mozambique, for the Delta and Omicron waves, the final *S_i_* estimated range of the prior wave (Alpha and Delta, respectively) was used as the susceptible initial prior, e.g. the *final S_i_* range of Alpha wave = *initial prior S_i_* range of Delta wave (see Table S5). In Zimbabwe, because data were only available for the Omicron (BA.1) wave, *initial prior S_i_* was set to that of Mozambique for the same outbreak, as both countries are in the same region and experienced similar epidemic patterns.

The depletion of susceptibles, *S_i_* at the end of an outbreak, reflected an approximation of the total new infections (both reported and unreported), given ongoing filter adjustment. Consequently, to estimate the cumulative disease burden from the beginning of the pandemic across subsequent waves, computation was carried out as follows:

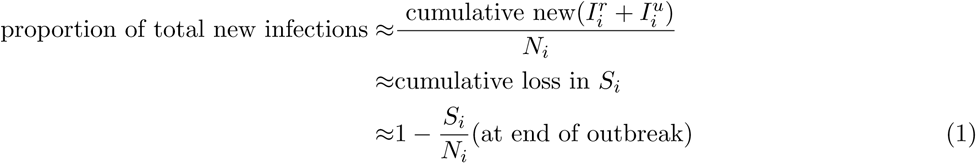

### 2.2 The SEIR-EAKF framework

The EAKF assimilation algorithm has been used in previous studies of infectious disease dynamics to assimilate observations and conduct inference [10, 33, 37]. When coupled with the dynamic model, Equation (S1)–Equation (S4), the combined SEIR-EAKF system iteratively optimizes the distribution of model state variables and parameters whenever new observations become available [3].

In general, sequential ensemble filtering is the problem of estimating the probability of the system state at a given time **x***_t_* conditional on observations *O_t_*taken up until and inclusive of time *t*. After model initialization, the system integrates an ensemble of simulations forward in time to compute the prior distribution for the model state variables, parameters, and the model-simulated case data. At the time of observation, the system is halted and the EAKF is used to update the state variables and parameters based on those model-generated prior estimates and case data. The EAKF algorithm is applied with prescribed observational error variance (OEV), see Section 2.2.3. In this fashion, the ensemble simulations of the observed state variables (incidence) are updated to align with observations (see Anderson [3] for algorithmic details). The updates (posteriors) are determined by computing the Kalman gain using the latest observation and the distribution of current model states (the prior). The unobserved state variables and model parameters are then adjusted by the EAKF using cross ensemble co-variability. The process is then repeated, with the posterior integrated to the next observation. Through this iterative optimization process, the ensemble of model simulations is better aligned to simulate current outbreak dynamics and estimate key parameters.

Here, a 300-member ensemble of simulations using the SEIR model Equation (S1)–Equation (S4) was coupled with the EAKF and case data. The state vector at time *t* is: **x***_t_* = (**s***_t_, θ_t_*), where **s***_t_* = [*S_i_, E_i_, I_i_^r^, I_i_^u^*] is the vector of the local state variables at time *t* and *θ_t_* = [*µ, Z, D, β_i_, α_i_*] is the vector of model parameters. Using observation *O_t_^i^* at time *t*, the posterior distribution of the system state is derived by applying Bayes’ rule (posterior ∝ prior *×* likelihood) to incorporate the new information. The EAKF deterministically adjusts the prior distribution to a posterior using Bayes rule while assuming a Gaussian distribution for both the prior and likelihood, which allows estimation of the first two moments (mean and covariance), leaving the higher-order moments unchanged. Bayes’ rule provides a target for updating the system state given an observation:

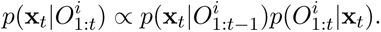

We used the daily number of new reported cases in province *i* on a given day *t*, *O_t_^i^*, as observations. Unobserved variables, such as the susceptible population *S*, and model parameters, were adjusted in accordance with covariant relationships with the observed variable i.e. reported new infections, 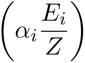 that arise naturally as a result of the dynamics of the system (see Anderson [3] for algorithmic details).

#### 2.2.1 State variables and parameter updates

State variables and the inferred model parameters (*β_i_* and *α_i_*) were updated locally for each province. Estimating the parameters locally was motivated by the discrepancies found between population size and total incidence: Nampula, the most densely populated province in Mozambique, reported the fewest COVID-19 cases, whereas Maputo Cidade, the least populated province, reported the highest number of cases across all variant waves. This discrepancy may be due to the limited healthcare accessibility in Nampula and lower population density compared to Maputo Cidade, which has the highest level of healthcare access and greatest population density among Mozambique provinces [12]. Similarly, for Zimbabwe, Harare province is the most densely populated province with better healthcare facilities compared to other provinces. Such differences underpinned an expectation of different contact and ascertainment rates and motivated deriving local *β_i_* and *α_i_* estimates for both countries.

The EAKF prediction–update cycle is performed sequentially, and an update is triggered by the arrival of new data (daily in this study) [3]. For the observable state variables, the *i^th^*ensemble member is updated by Equation (S13), while, for the unobserved variables/parameters *x^i^*, the *i^th^* ensemble member is updated by Equation (S14).

#### 2.2.2 Initial priors

The initial 300-member ensemble of state variables and parameters were randomly drawn from a uniform distribution using Latin hypercube sampling and the initial range of values presented in Table S5; other parameters, *D*, *Z*, and *µ* were fixed. During filtering, ensemble members could move outside initial ranges; in such instances, the individual ensemble member would be resampled from the prior range.

The initial prior range of *α_i_* = [0.004 *−* 0.1] (which is low, corresponding to a maximum of one reported case for every ten infected) was motivated by the study of Han *et al.* [18], which estimated an overall daily report rate (ascertainment rate) ranging from *α* = [0.0002 *−* 0.3], among all African countries. Furthermore, the study by Evans *et al.* [17] highlighted that the lower-than-expected case burdens in Madagascar were explained solely by detection rates of 0.1–1% (or *α* = [0.001 *−* 0.01]). On the other hand, due to the the heterogeneity of reported cases across provinces in Mozambique, the initial prior lower bound for *α_i_* was varied by province for both countries (see Table S5). For province *i*, we applied the formula: lower bound *α_i_* = (total reported cases)*/*0.5*N_i_*, assuming 50% of the population was infected at the end of the three variant waves (Alpha, Delta, Omicron (BA.1)). An exception was Maputo Cidade which had a different initial prior range *α_i_*= [0.14, 0.2], because, despite being the less populous province it reported the highest number of cases. This may be attributed to Maputo Cidade being the capital city with greater access to healthcare and testing facilities compared to the rest of the country. Generally, the remaining Mozambique provinces had lower bounds ranging *α_i_* = [0.004 *−* 0.03], whereas for Zimbabwe, lower bounds were *α_i_* = [0.01 *−* 0.04]. The *S_i_* initial priors varied for each wave because the model was run piecewise for the periods as described above and in Table S5. Due to missing Zimbabwe COVID-19 provincial case data for earlier waves, the initial S prior for the Omicron (BA.1) variant wave was based on that of Mozambique.

#### 2.2.3 Observational error variance

Using the EAKF requires specification of error for both the simulated model output and observations. The model error may be easily computed as the variance of the 300-member ensemble. For each *O^i^*, we specify the OEV at time *t* as:

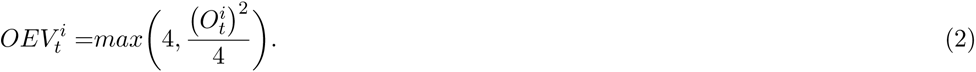

An OEV of this form has been used successfully for inference and forecasting for infectious diseases such as influenza [47, 33] and COVID-19 [22].

#### 2.2.4 Filter divergence

As successive observations are assimilated, there is a tendency for the variance between the ensemble members to decrease due to repeated filter adjustment. This may potentially lead to filter divergence, in which the ensemble error variance is so minimal that the fitting process essentially ignores the observations [3]. To prevent filter divergence, the prior ensemble was inflated by a multiplicative factor *λ* = 0.015, before each daily assimilation and calculation of the posterior (see Equation (S15)). The inflation was applied to all state variables and estimated parameters, i.e. *β_i_* (transmission rate) and *α_i_* (ascertainment rate).

### 2.3 Synthetic Testing

To validate EAKF inference with the SEIR model, we generated a synthetic, model-simulated COVID-19 outbreak (defined as the ’truth’), defined by model Equation (S1)–Equation (S4) and we tested the ability of the model/filter framework to identify the true model state variables and parameters . Specifically, we generated nine synthetic datasets using different scenarios combinations of transmission rates, *β_i_* = (0.8, 1.3, 1.8) and ascertainment rates, *α_i_* = (0.01, 0.04, 0.09), where the chosen parameter values were the same across all provinces for each outbreak, as shown in Table S6. Synthetic observations of new daily reported cases (reported new infections, 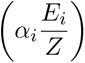 were then generated by adding normally distributed random observational error calculated as described in Equation (2). The resulting time series of synthetic error-laden observational records were smoothed using a 7-day moving average, and then used for assimilation in the combined SEIR-EAKF framework to determine whether the system can reliably identify known parameter values.

### 2.4 Parameter estimation using real data

In order to estimate the transmission rate, *β_i_* and ascertainment rate, *α_i_*, for each variant wave (Alpha, Delta and Omicron (BA.1)), we fitted the model to province specific case data (i.e., number of new reported cases) as reported by Mozambique Ministry of Health-SIGILIA and DISA [29] and Zimbabwe COVID-19 hub [1] for the period between March 2020 to December 2022. Due to missing Zimbabwe COVID-19 provincial data we only estimated parameters for the Omicron (BA.1) variant wave in Zimbabwe.

The SEIR-EAKF framework was applied in isolation to the eleven Mozambique provinces and, separately, the ten Zimbabwe provinces, as illustrated in Figure S1. The Mozambique COVID-19 data was transformed from weekly cases to daily cases using a linear interpolation method, and piecewise parameter estimation was carried out for the model’s local parameters, *β_i_* and *α_i_*, for each variant wave as described in Table S4 and Section S1.7.

## 3 Results

### 3.1 Data description

#### COVID-19 data

The reported daily COVID-19 cases used for this study span three variant waves (Alpha, Delta, and Omicron (BA.1)) during the first three years of the pandemic, i.e. from March 30*^th^* 2020 to December 26*^th^* 2022 for Mozambique and from November 18*^th^* 2021 to January 26*^th^* 2022 for Zimbabwe. The available Zimbabwe provincial data began in the middle of the Delta variant wave, therefore, we only utilized the data covering the Omicron (BA.1) wave. At the provincial scale, COVID-19 daily cases were obtained from the Zimbabwe COVID-19 hub [1] and weekly cases for Mozambique were obtained from the Mozambique Ministry of Health-SIGILIA and DISA [29]. Heat maps of daily and weekly counts of COVID-19 cases, for each variant period, in each province of both countries are presented in Figure 2 while plots of the disaggregated time series of COVID-19 infections for the provinces are reported in Figure S1.

**Figure 2:**
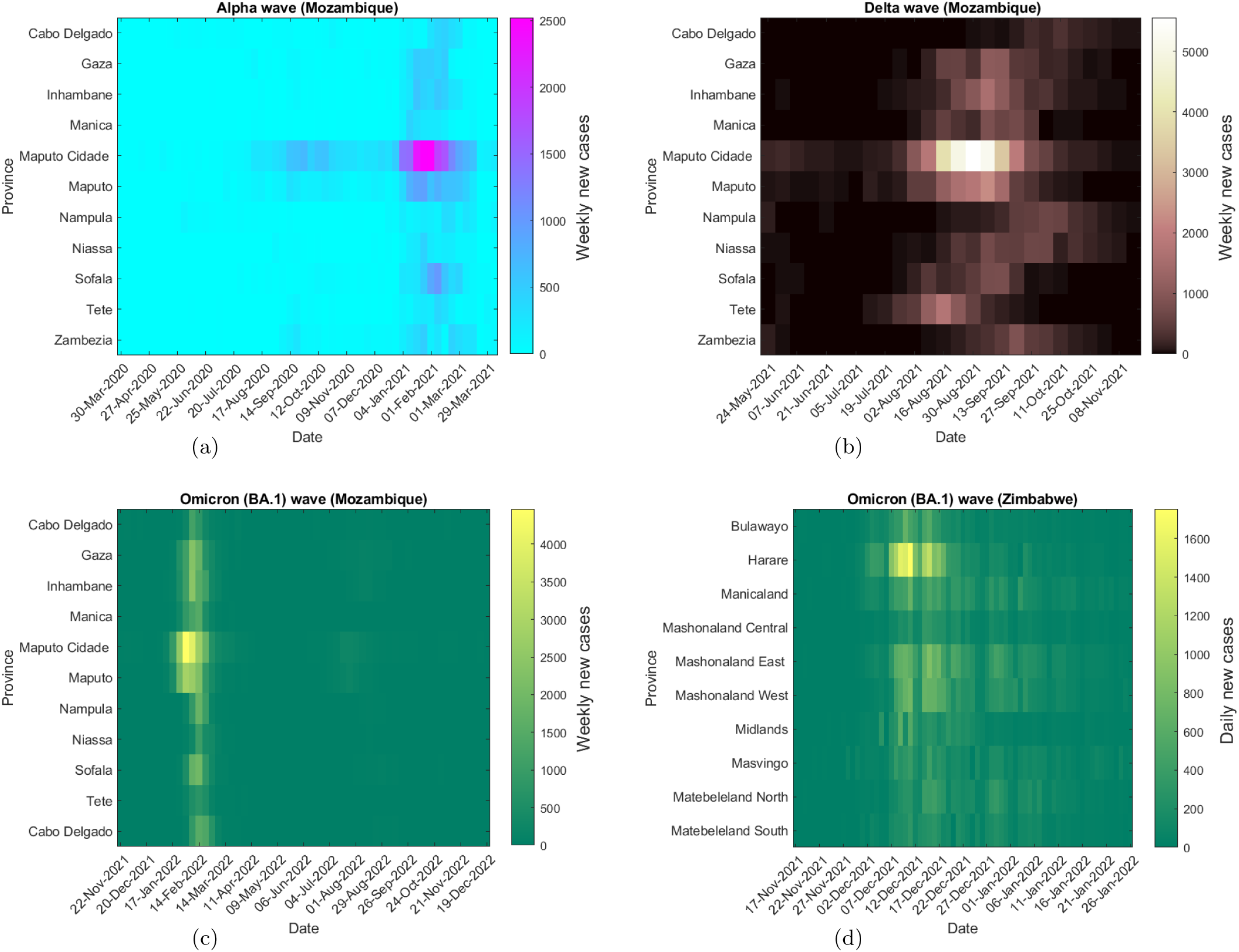
Heat maps of daily and weekly provincial level COVID-19 infections in Mozambique and Zimbabwe between March 2020 and December 2022, based on data released by the Mozambique Ministry of Health-SIGILIA and DISA [29] and Zimbabwe COVID-19 hub [1].

#### Population data

Provincial population data were taken from the Zimbabwe National Statistics Agency [2] and Instituto Nacional de Estatistica Moçambique [9] for Zimbabwe and Mozambique, respectively.

### 3.2 Validation of the model-inference system using synthetic outbreaks

The synthetic observations of COVID-19 cases, along with their defined OEV, were used to test whether the SEIR-EAKF system could estimate unobserved state variables and parameters (*β_i_* and *α_i_*) accurately. Creffig:paramsSynth depicts the parameter estimates for the simulation period; the posterior parameters estimates (*β_i_* and *α_i_*) converged near their target values and the reproductive number at the end of the variant wave, *Rt_i_* was correctly estimated. The ensemble posterior distributions at the end of the outbreak were well constrained for all the generated outbreaks as shown in Figure S3. The results indicate that an ensemble of size 300 was sufficient to capture the target ’true’ parameters.

### 3.3 Mozambique parameter estimates

The estimates for *β_i_* and *α_i_* at the end of each variant outbreak, along with their associated 95% credible intervals (CrIs) are presented in Table 1. With the exception of Maputo Cidade, for all provinces, the mean estimates across all three waves were in the range *α_i_* = [0.08 *−* 0.09]. On the other hand, for Maputo Cidade, with its unique *α_i_* prior range of [0.14, 0.2] (see Section S1.7), the estimate was around *α_i_* = 0.17 across all variant waves. The local *β_i_* mean estimates and by extension *Rt_i_* increased with each successive wave: the mean estimate ranges of *β_i_* were [0.6*−*0.7] (*Rt_i_* = [1.4 *−* 1.8]), for the Alpha wave, slightly higher, *β_i_* = [0.6 *−* 1.4] (*Rt_i_* = [1.4 *−* 4]), for the Delta wave, and highest for the Omicron (BA.1) wave, *β_i_*= [1.1 *−* 1.7] (*Rt_i_*= [3 *−* 5]).

**Table 1:** EAKF estimated parameters and credible intervals at the end of each Mozambique variant period.

| Province | Parameter | Prior | EAKF estimate (Alpha) |  | EAKF estimate (Delta) |  | EAKF estimate (Omicron (BA.1)) |  |
| --- | --- | --- | --- | --- | --- | --- | --- | --- |
|  |  |  | mean | 95% CrI | mean | 95% CrI | mean | 95% CrI |
| transmission rates |  |  |  |  |  |  |  |  |
| Cabo Delgado | $\beta_1$ | [0.5, 2.0] | 0.6038 | (0.5032, 0.8037) | 1.1571 | (0.7873, 1.6191) | 1.1528 | (1.0684, 1.4876) |
| Gaza | $\beta_2$ | | 0.5643 | (0.5036, 0.7021) | 1.4886 | (0.6507, 1.9467) | 1.5630 | (0.6812, 1.9322) |
| Inhambane | $\beta_3$ | | 0.5830 | (0.5040, 0.7898) | 1.2970 | (0.6593, 1.9414) | 1.6998 | (0.9363, 1.9593) |
| Manica | $\beta_4$ | | 0.5526 | (0.5021, 0.6768) | 0.6362 | (0.5073, 1.3342) | 1.4411 | (0.6655, 1.9614) |
| Maputo Cidade | $\beta_5$ | | 0.6681 | (0.5157, 0.9132) | 1.1154 | (0.5938, 1.9074) | 1.5664 | (0.7519, 1.9561) |
| Maputo | $\beta_6$ | | 0.6136 | (0.5135, 0.8306) | 1.3520 | (0.6281, 1.9099) | 1.5299 | (0.8293, 1.9683) |
| Nampula | $\beta_7$ | | 0.5645 | (0.5035, 0.6679) | 0.9902 | (0.6117, 1.7501) | 1.1174 | (0.5324, 1.9316) |
| Niassa | $\beta_8$ | | 0.6059 | (0.5026, 0.7684) | 0.7763 | (0.5156, 1.5475) | 1.5890 | (0.7134, 1.9788) |
| Sofala | $\beta_9$ | | 0.6060 | (0.5046, 0.8032) | 0.5680 | (0.5035, 0.9613) | 1.3762 | (0.7603, 1.9652) |
| Tete | $\beta_{10}$ | | 0.6082 | (0.5036, 0.8303) | 0.6315 | (0.5107, 1.1309) | 1.1819 | (0.5908, 1.9180) |
| Zambezia | $\beta_{11}$ | | 0.5796 | (0.5055, 0.7656) | 0.8914 | (0.5229, 1.7148) | 1.3361 | (0.6657, 1.9511) |
| ascertainment rates |  |  |  |  |  |  |  |  |
| Cabo Delgado | $\alpha_1$ | [0.007, 0.1] | 0.0923 | (0.0744, 0.0993) | 0.0833 | (0.0158, 0.0990) | 0.0798 | (0.0301, 0.0983) |
| Gaza | $\alpha_2$ | [0.02, 0.1] | 0.0883 | (0.0606, 0.0998) | 0.0622 | (0.0264, 0.0978) | 0.0718 | (0.0316, 0.0991) |
| Inhambane | $\alpha_3$ | [0.03, 0.1] | 0.0904 | (0.0554, 0.0998) | 0.0627 | (0.0330, 0.0965) | 0.0881 | (0.0570, 0.0991) |
| Manica | $\alpha_4$ | [0.01, 0.1] | 0.0922 | (0.0739, 0.0996) | 0.0881 | (0.0426, 0.0987) | 0.0836 | (0.0203, 0.0994) |
| Maputo Cidade | $\alpha_5$ | [0.14, 0.2] | 0.1739 | (0.1419, 0.1988) | 0.1760 | (0.1477, 0.1990) | 0.1729 | (0.1411, 0.1977) |
| Maputo | $\alpha_6$ | [0.03, 0.1] | 0.0836 | (0.0558, 0.0985) | 0.0640 | (0.0334, 0.0986) | 0.0677 | (0.0379, 0.0970) |
| Nampula | $\alpha_7$ | [0.004, 0.1] | 0.0911 | (0.0658, 0.0995) | 0.0318 | (0.0174, 0.0754) | 0.0867 | (0.0163, 0.0987) |
| Niassa | $\alpha_8$ | [0.01, 0.1] | 0.0930 | (0.0684, 0.0998) | 0.0812 | (0.0371, 0.0992) | 0.0888 | (0.0403, 0.0979) |
| Sofala | $\alpha_9$ | [0.01, 0.1] | 0.0872 | (0.0519, 0.0997) | 0.0849 | (0.0455, 0.0995) | 0.0827 | (0.0176, 0.0998) |
| Tete | $\alpha_{10}$ | [0.009, 0.1] | 0.0902 | (0.0684, 0.0995) | 0.0862 | (0.0468, 0.0996) | 0.0787 | (0.0468, 0.0996) |
| Zambezia | $\alpha_{11}$ | [0.005, 0.1] | 0.0902 | (0.0543, 0.0996) | 0.0884 | (0.0320, 0.0990) | 0.0879 | (0.0513, 0.0996) |
| reproductive number |  |  |  |  |  |  |  |  |
| Cabo Delgado | $Rt_1$ | | 1.5367 | (1.2760, 2.0486) | 2.9193 | (1.9882, 4.0874) | 2.9056 | (1.3940, 4.8801) |
| Gaza | $Rt_2$ | | 1.4317 | (1.2738, 1.7767) | 3.6994 | (1.6392, 4.8517) | 3.9199 | (1.7308, 4.8177) |
| Inhambane | $R_3$ | | 1.4748 | (1.2827, 2.0140) | 3.2424 | (1.6336, 4.8583) | 4.3049 | (2.3873, 4.9700) |
| Manica | $Rt_4$ | | 1.4100 | (1.2765, 1.7275) | 1.6184 | (1.2840, 3.3278) | 3.6355 | (1.6712, 4.9141) |
| Maputo Cidade | $Rt_5$ | | 1.7843 | (1.3755, 2.4536) | 2.9981 | (1.5922, 5.1157) | 4.1683 | (1.9782, 5.2456) |
| Maputo | $Rt_6$ | | 1.5550 | (1.3017, 2.0922) | 3.3779 | (1.5619, 4.8312) | 3.8328 | (2.1190, 4.9489) |
| Nampula | $Rt_7$ | | 1.4404 | (1.2798, 1.7002) | 2.4349 | (1.5105, 4.2809) | 2.7798 | (1.3296, 4.8345) |
| Niassa | $Rt_8$ | | 1.5362 | (1.2773, 1.9420) | 1.9631 | (1.3020, 3.8910) | 4.0304 | (1.8178, 4.9980) |
| Sofala | $Rt_9$ | | 1.5218 | (1.2829, 2.0447) | 1.4373 | (1.2688, 2.4461) | 3.4886 | (1.9383, 4.9815) |
| Tete | $Rt_{10}$ | | 1.5395 | (1.2799, 2.1200) | 1.5965 | (1.2932, 2.8883) | 2.9824 | (1.5107, 4.8073) |
| Zambezia | $Rt_{11}$ | | 1.4710 | (1.2825, 1.9245) | 2.2548 | (1.3290, 4.3444) | 3.4098 | (1.6867, 4.9622) |

Figure S4–Figure S6 illustrate the time evolution of the parameter posteriors (means and 95% CrIs) for the Alpha, Delta and Omicron (BA.1) variant waves, respectively, plotted against their prior range. The initial drift in the first month, is likely a reflection of the time needed for the EAKF to learn the system [3]. The *α_i_* estimates at the end of the outbreaks were closer to the upper bound of 0.2 for Maputo Cidade and 0.1 for the rest of the provinces. For the Alpha and Delta waves *β_i_* converges to a solution during January 2021 and August 2021, respectively, as cases rise, on the other hand, for the Omicron (BA.1) wave, *β_i_*convergence to a solution is less clear, with a broader posterior and higher mean estimate.

The posterior fits of new reported cases to province-specific actual data [29] were generally good for all provinces in Mozambique for all variant waves, as demonstrated in Figure S7–Figure S9. The model-inference framework was able to reproduce even complicated outbreak structures characterized by multiple peaks such as during the Alpha wave in Nampula, Tete and Zambezia.

In order to check the consistency of the parameter estimates for each variant wave, presented in Table 1, SEIR-EAKF system simulations were repeated 20 times. The mean parameter estimates (*β_i_*and *α_i_*), and the corresponding mean *Rt_i_*for the runs are illustrated in Figure 3 and show consistency for most estimates. Figure S12 (a)-(c) shows the posterior mean estimates of susceptibility % at the end of each variant wave. An estimated 75 *−* 95% of the population remained susceptible at the end of the Alpha wave (in March 2021), furthermore, 15 *−* 50% and 5 *−* 30% remained susceptible after the Delta (in October 2021) and Omicron (BA.1) (in March 2022) waves, respectively.

**Figure 3:**
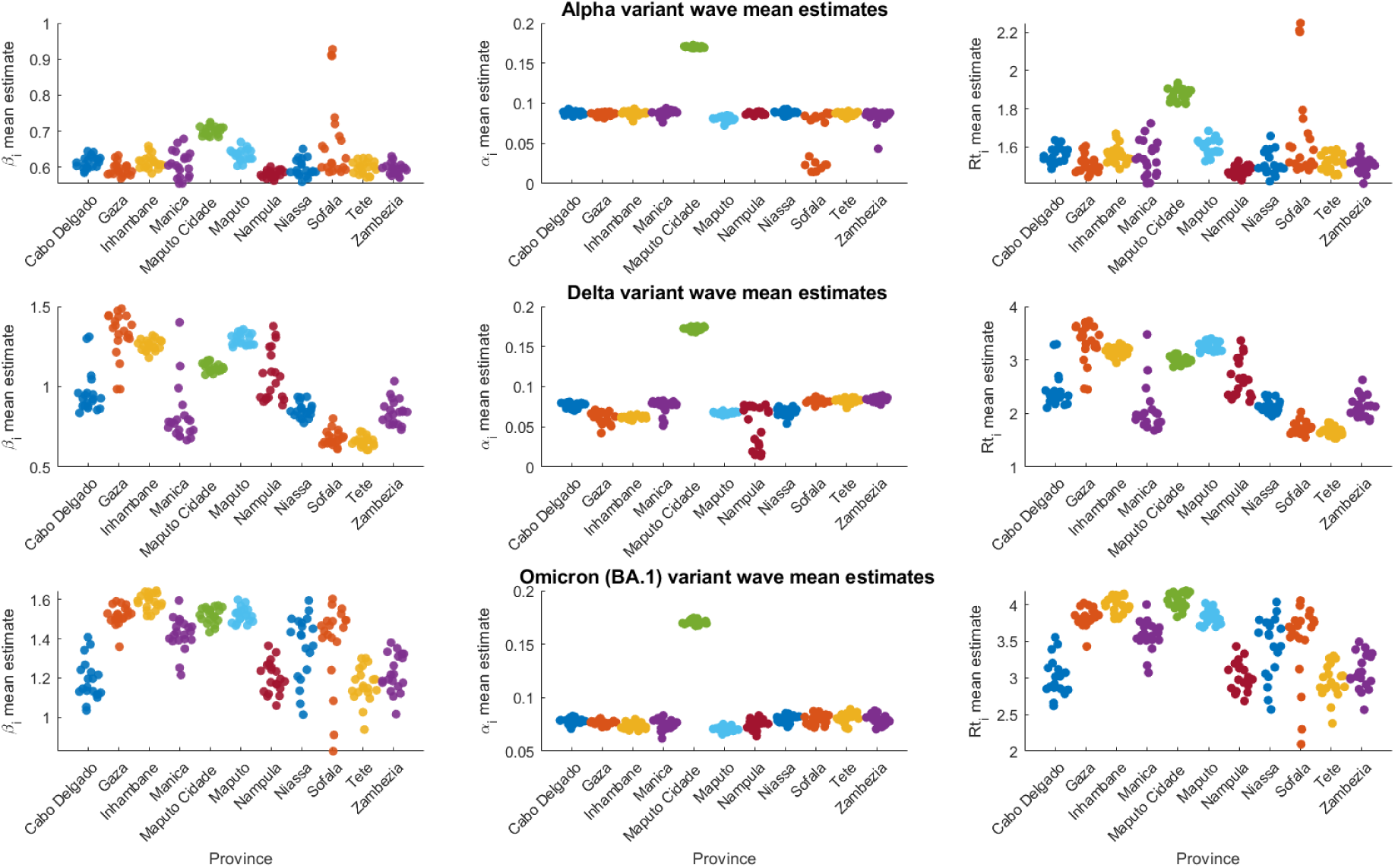
Distribution of mean parameter estimates (*α_i_*and *β_i_*, and the corresponding *Rt_i_*) for the 11 Mozambique provinces at the end of variant waves (Alpha, Delta and Omicron (BA.1)) for 20 runs.

To quantify the burden of COVID-19 infections at the end of each variant period, we estimated the total cumulative infections, i.e., both reported and unreported infections by applying Equation (1). Figure 4(a)–Figure 4(c) display the spatial distribution of the estimated total new infections (reported and unreported) across all Mozambique provinces. We estimated an increase in the proportion of the infected population across the three VOC (Alpha, Delta and Omicron (BA.1)): the Alpha wave estimates were below 20% for the majority of the country; in contrast, Maputo Cidade and Maputo, showed the highest burdens of 72% and 57%, respectively. The cumulative infection estimates increased from 6 *−* 72% (Alpha wave), to 46 *−* 88% (Delta wave), to 74 *−* 95% (Omicron BA.1 wave).

**Figure 4:**
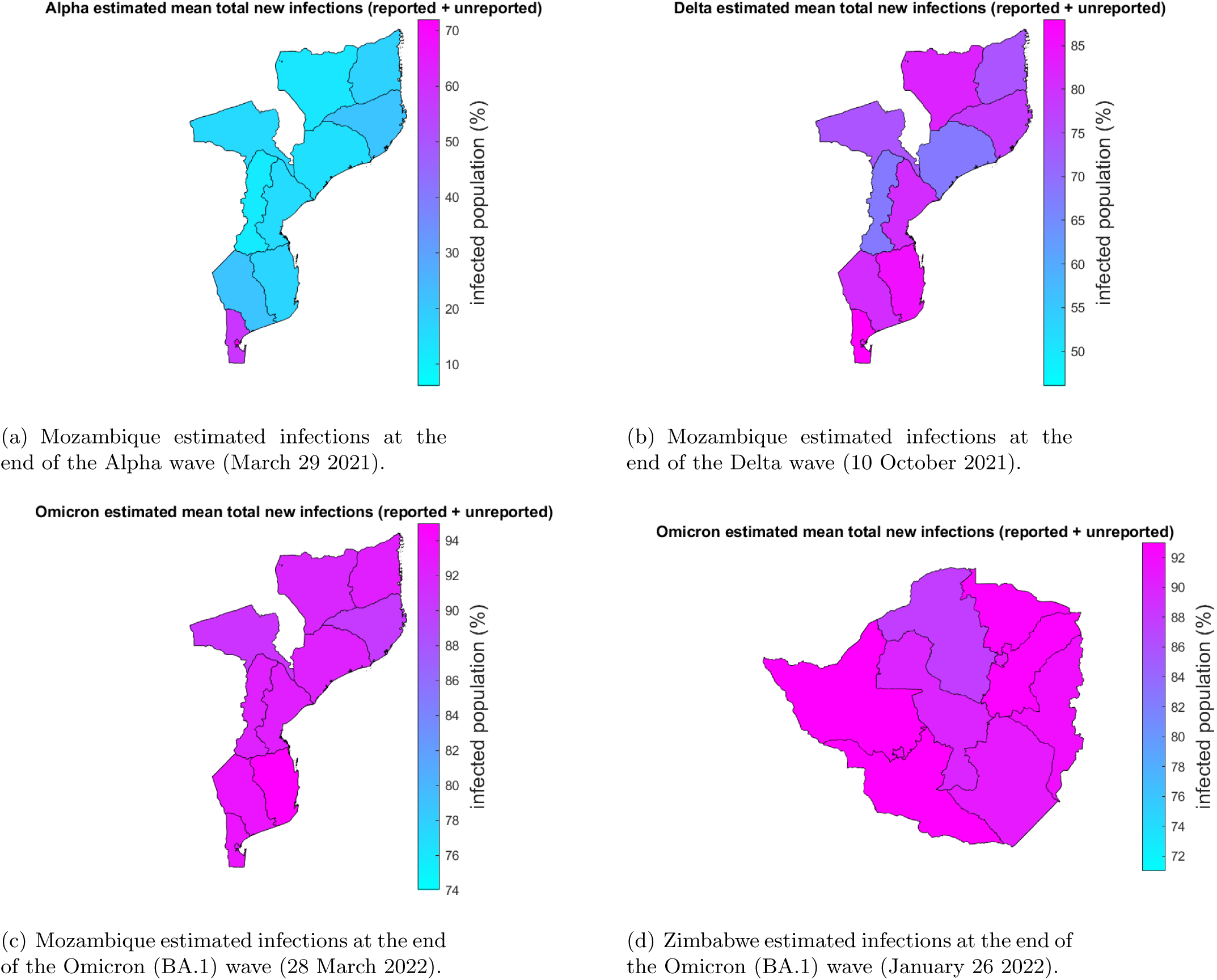
Spatial distribution of estimated SARS-CoV-2 infections (both reported and unreported) per VOC for Mozambique and Zimbabwe provinces between March 2020 and December 2022 [29, 1]. Colors toward red indicate more infections.

### 3.4 Zimbabwe parameter estimates

Table 2 presents the parameters estimates (*β_i_* and *α_i_*) and associated 95% CrIs at the end of the Omicron (BA.1) variant outbreak. The estimates for the ascertainment rate were in the range *α_i_* = [0.08 *−* 0.09], which is in line with Mozambique estimates (shown in Table 1). On the other hand, the mean estimates for the transmission rate ranged from *β_i_* = [1.1 *−* 1.7] for all provinces with a reproductive number of *Rt_i_* = [3 *−* 5]. The time evolution of *α_i_* and *β_i_* parameter posteriors (means and 95% CrI) over the course of the Omicron (BA.1) wave, plotted against their prior range is demonstrated in Figure S10. The *α_i_* estimates drifted higher towards the upper bound (0.1) during December 2021, a level maintained until the end of the outbreak. The convergence of *β_i_* to a solution is less clear, with a broader posterior and higher mean estimate.

**Table 2:** EAKF estimated parameters and credible intervals for the Zimbabwe Omicron (BA.1) variant period.

| Province | Parameter | Prior | EAKF estimate (Omicron (BA.1)) |  |
| --- | --- | --- | --- | --- |
|  |  |  | mean | 95% CrI |
| transmission rates |  |  |  |  |
| Bulawayo | $\beta_1$ | [0.5, 2.0] | 1.6529 | (1.0081, 1.9813) |
| Harare | $\beta_2$ | | 1.6407 | (1.0550, 1.9762) |
| Manicaland | $\beta_3$ | | 1.1327 | (0.5971, 1.9040) |
| Mashonaland Central | $\beta_4$ | | 1.2506 | (0.7189, 1.9148) |
| Mashonaland East | $\beta_5$ | | 1.5438 | (0.8128, 1.9457) |
| Mashonaland West | $\beta_6$ | | 1.4040 | (0.8200, 1.9323) |
| Midlands | $\beta_7$ | | 1.3669 | (0.6988, 1.9702) |
| Masvingo | $\beta_8$ | | 1.5457 | (0.8012, 1.9652) |
| Matebeleland North | $\beta_9$ | | 1.6567 | (1.0512, 1.9716) |
| Matebeleland South | $\beta_{10}$ | | 1.6446 | (0.9344, 1.9848) |
| ascertainment rates |  |  |  |  |
| Bulawayo | $\alpha_1$ | [0.04, 0.1] | 0.0911 | (0.0668, 0.0997) |
| Harare | $\alpha_2$ | [0.03, 0.1] | 0.0924 | (0.0744, 0.0997) |
| Manicaland | $\alpha_3$ | [0.02, 0.1] | 0.0864 | (0.0387, 0.0990) |
| Mashonaland Central | $\alpha_4$ | [0.01, 0.1] | 0.0929 | (0.0589, 0.0997) |
| Mashonaland East | $\alpha_5$ | [0.03, 0.1] | 0.0871 | (0.0562, 0.0984) |
| Mashonaland West | $\alpha_6$ | [0.02, 0.1] | 0.0930 | (0.0694, 0.0997) |
| Midlands | $\alpha_7$ | [0.01, 0.1] | 0.0917 | (0.0534, 0.0993) |
| Masvingo | $\alpha_8$ | [0.02, 0.1] | 0.0876 | (0.0474, 0.0989) |
| Matebeleland North | $\alpha_9$ | [0.04, 0.1] | 0.0857 | (0.0499, 0.0992) |
| Matebeleland South | $\alpha_{10}$ | [0.03, 0.1] | 0.0846 | (0.0489, 0.0991) |
| reproductive number |  |  |  |  |
| Bulawayo | $Rt_1$ | | 5.4010 | (3.3035, 6.4817) |
| Harare | $Rt_2$ | | 5.3709 | (3.4402, 6.4669) |
| Manicaland | $Rt_3$ | | 3.6866 | (1.9242, 6.2350) |
| Mashonaland Central | $Rt_4$ | | 4.0797 | (2.3522, 6.2600) |
| Mashonaland East | $Rt_5$ | | 5.0435 | (2.6502, 6.3627) |
| Mashonaland West | $Rt_6$ | | 4.6015 | (2.6877, 6.3179) |
| Midlands | $Rt_7$ | | 4.4749 | (2.2919, 6.4159) |
| Masvingo | $Rt_8$ | | 5.0560 | (2.0240, 3.0989) |
| Matebeleland North | $Rt_9$ | | 5.4064 | (3.4283, 6.4578) |
| Matebeleland South | $Rt_{10}$ | | 5.3699 | (3.0497, 6.4785) |

Figure S11 demonstrates a good model fit to provincial data [1] for the outbreak with the posterior estimates capturing the outbreak peaks for all ten provinces. The consistency of the parameter estimates Table 2 was evaluated by running the model 20 times, and the distribution of the mean parameter estimates (*β_i_* and *α_i_*) at the end of the outbreak, and the corresponding mean *Rt_i_* for the runs are shown in Figure 5. Figure S12 (d) shows the posterior mean estimates of susceptibility % at the end of the Omicron (BA.1) wave (in January 2022), where 15 *−* 40% of the population remained susceptible at the end of that outbreak.

**Figure 5:**
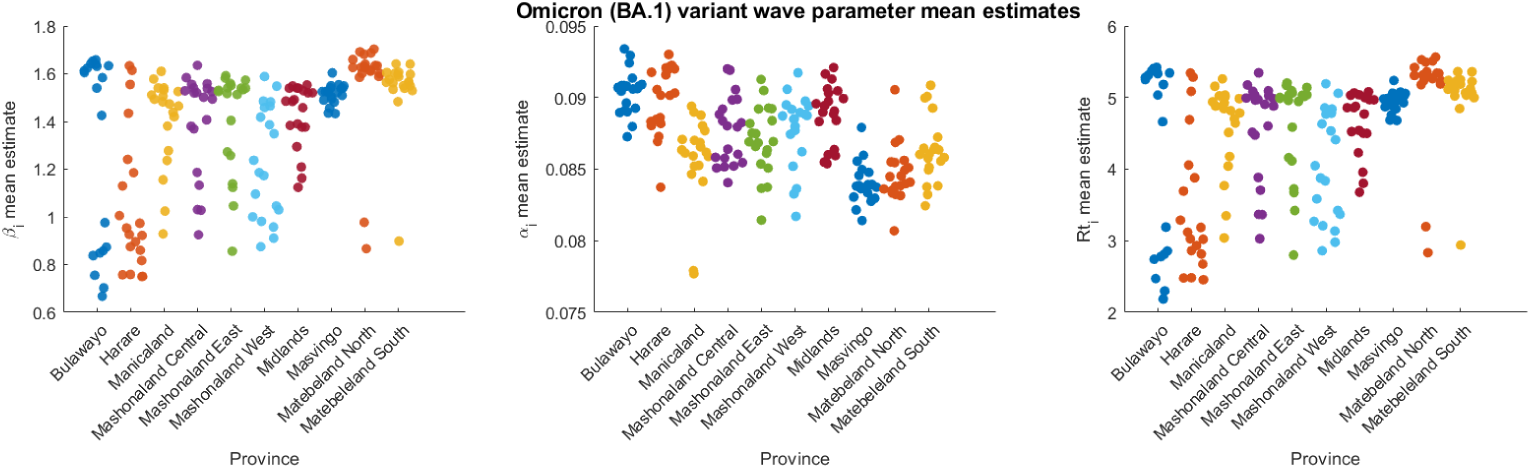
Distribution of mean parameter estimates (*α_i_*, *β_i_*, and corresponding *Rt_i_*) for th 10 Zimbabwe provinces at the end of Omicron (BA.1) wave for 20 runs.

The spatial distribution of the estimated mean total new COVID-19 infections (both reported and unreported) at the end of the Omicron (BA.1) wave is summarized in Figure 4(d). The findings indicate that 71 *−* 93% of the population had been infected.

## 4 Discussion

Although the emergence of COVID-19 and its rapid spread created a public health emergency of international concern, the impact of the pandemic in Sub-Saharan Africa, as documented in cases, hospitalizations and deaths, appears far lower than in the Americas, Europe, and Asia [43]. We utilized a model-inference framework to shed light on disease dynamics in two southern African countries, i.e., Mozambique and Zimbabwe, during the first three years of the pandemic. While accounting for undocumented cases, we estimated key epidemiologic parameters namely the transmission rate, and the ascertainment rate at the end of each outbreak. Our study provides several insights into the disease burden of infections (reported and unreported) for three VOCs, i.e. Alpha, Delta and Omicron (BA.1). While we focus on the Mozambique and Zimbabwe case, our framework can be applied to other African countries given the low documented impact of the disease in this region.

The ascertainment rate, *α*, estimates were very low for Mozambique and Zimbabwe provinces, matching our prior assumption. Our estimates in both countries is in line with the studies by Han *et al.* [18] and Evans *et al.*[17]. This indicates that the majority of infections were never documented as cases; by extension, many severe cases requiring hospitalization or resulting in death may have been missed due to limited access to healthcare and testing facilities. The estimated low ascertainment rate highlights the need for enhanced reporting and surveillance mechanisms, with special emphasis on Sub-Saharan Africa. Mozambique experienced natural disasters and conflict which hampered access to healthcare facilities: Prior to the pandemic, the country was devastated by two consecutive deadly and destructive cyclones (Idai and Kenneth), which caused massive destruction to infrastructure; consequently, accessibility in some places became more difficult. Furthermore, conflict in northern Mozambique produced additional challenges during the pandemic by displacing many people, hence making it difficult to control the disease [26].

The transmission rate estimates, and by extension reproductive number, rose with each successive wave. This matches observations in the Americas, Africa, Europe and Asia that the Omicron variant, in particular, was more transmissible than the preceding Delta variant [21, 24], which in turn had greater transmissibility than the preceding Alpha variant [15, 27]. Mozambique and Zimbabwe deployed public health and social measures to mitigate the spread of COVID-19. These included travel restrictions, lockdown, social distancing measures, compulsory mask wearing, contact tracing and testing, school closures and use of personal protective equipment among health workers [4, 5, 13, 14].

Our model was fitted to provincial reported cases. We find that representing and accounting for unreported cases across provinces is crucial for estimating the true disease burden, i.e. new infections (reported and unreported), at the end of each variant wave. The low numbers of confirmed cases in Africa were generally a poor indicator of true incidence of infection, which may be attributed to limited testing capacity and health system access. Overall, the estimated disease burden in Mozambique and Zimbabwe was much higher than the documented reported cases for each VOC (see [29, 1], Table S1, and Table S2). The estimates highlight that by the end of the Omicron (BA.1) wave, almost the entire population had been infected, which is in agreement with the estimates of 93% cumulative total infection after the same wave in South Africa [46]. Our results indicate that in African countries, efforts must be made to improve the timely reporting and surveillance of public health threats.

There are limitations to our model: Even though human mobility plays a crucial role in the transmission of SARS-CoV-2, the unavailability of inter-provincial human movement data for both Mozambique and Zimbabwe was a drawback. We explored utilizing a gravity model to estimate human movement between the provinces. This approach, however, did not yield credible results; hence, we implemented the metapopulation model without movement. Additionally, there was limited provincial case data for Zimbabwe, which covered only the Omicron (BA.1) variant. Furthermore, the model does not explicitly account for re-infections, but the filter adjusts the susceptibles, *S_i_*, thereby implicitly accounting for increasing susceptibility due to immune escape.

In conclusion, we developed an inference-based transmission model to aid in understanding the evolving dynamics of SARS-CoV-2 in Mozambique and Zimbabwe. By taking unreported cases into account, we estimated key epidemiologic characteristics, i.e. the transmission rate and ascertainment rate. This approach and findings are relevant for countries with less comprehensive surveillance systems. The findings of this study on the disease burden can help guide future public health planning. In particular, they shed light on respiratory virus transmission dynamics in two African countries little investigated to date.

## Data Availability

The data that supports the findings of this study are available upon request from INS Mozambique [Sergio Chicumbe -Directorate for health research,

