## supplemental material for "SARS-CoV-2 transmission dynamics in Mozambique and Zimbabwe during the First Three Years of the Pandemic"

#### S1 Methodology

##### S1.1 Disease dynamics

Table S1: State variables and parameters of the model

| **State variables** | **Description** | **Units** |
|---|---|---|
| $S_i$ | susceptible population in province $i$ | *individuals* |
| $E_i$ | exposed population in province $i$ | *individuals* |
| $I_i^r$ | infected reported population in province $i$ | *individuals* |
| $I_i^u$ | infected unreported population in province $i$ | *individuals* |
| $R_i$ | recovered population in province $i$ | *individuals* |
| $N_i$ | total population in province $i$ | *individuals* |
| **Parameters** | **Description** | **Units** |
| $\beta_i$ | transmission rate in province $i$ | *1/day* |
| $\mu$ | reduction of infection rate for unreported infected individuals | *dimensionless* |
| $Z$ | average latency period | *days* |
| $\alpha_i$ | ascertainment rate in province $i$ | *dimensionless* |
| $D$ | average duration of infection | *days* |

Model disease dynamics are described by the following equations:

$$\frac{d}{dt}S_i = -\beta_i\frac{S_i I_i^r}{N_i} - \mu\beta_i\frac{S_i I_i^u}{N_i} \tag{S1}$$

$$\frac{d}{dt}E_i = \beta_i\frac{S_i I_i^r}{N_i} + \mu\beta_i\frac{S_i I_i^u}{N_i} - \frac{E_i}{Z} \tag{S2}$$

$$\frac{d}{dt}I_i^r = \alpha_i\frac{E_i}{Z} - \frac{I_i^r}{D} \tag{S3}$$

$$\frac{d}{dt}I_i^u = (1-\alpha_i)\frac{E_i}{Z} - \frac{I_i^u}{D} \tag{S4}$$

$$\frac{d}{dt}R_i = \frac{I_i^r}{D} + \frac{I_i^u}{D} \tag{S5}$$

$$N_i = S_i + E_i + I_i^r + I_i^u + R_i$$

Hence, for stochastic integration, equations (Equation (S1))–(Equation (S4)) were calculated as follows:

$$
\begin{aligned}
P1 =& Pois\left(\beta_i \frac{S_i I_i^r}{N_i}\right) \\
P2 =& Pois\left(\mu\beta_i \frac{S_i I_i^r}{N_i}\right) \\
P3 =& Pois\left(\alpha_i \frac{E_i}{Z}\right) \\
P4 =& Pois\left((1-\alpha_i) \frac{E_i}{Z}\right) \\
P5 =& Pois\left(\frac{I_i^r}{D}\right) \\
P6 =& Pois\left(\frac{I_i^u}{D}\right)
\end{aligned}
$$

$$\frac{d}{dt}S_i = -P1 - P2 \tag{S6}$$

$$\frac{d}{dt}E_i = P1 + P2 - P3 - P4 \tag{S7}$$

$$\frac{d}{dt}I_i^r = P3 - P5 \tag{S8}$$

$$\frac{d}{dt}I_i^u = P4 - P6 \tag{S9}$$

##### S1.2 Disease-free equillibrium, $\mathcal{E}_i$

The system of equations (Equation (S1))–(Equation (S4)) has three infected states, $E_i$, $I_i^r$, and $I_i^u$; and two uninfected states, $S_i$ and $R_i$ for each location $i$. Although five states exist in the model, it is four-dimensional as the total population size remains constant. At the disease-free steady state $E_i = I_i^r = I_i^u = R_i = 0$, hence $S_i = N_i$, and the disease-free equillibrium becomes $\mathcal{E}_i = (N_i, 0, 0, 0, 0)$.

##### S1.3 Reproductive number, $Rt_i$, for the locations

We calculated the reproductive number at a particular time, $Rt_i$ for each location using the model parameters. Specifically, $Rt_i$ is the largest eigenvalue of the next-generation matrix (NGM) [2, 10]. The linearization of equations (Equation (S2))–(Equation (S4)) is closed, in that it does not involve the deviation of S from its steady-state value, also, R does not appear in equations (Equation (S2))–(Equation (S4)). Hence, we have the linearized *infection subsystem*, which describes the production of new infections and changes in the states as:

$$\frac{d}{dt}E_i = \beta_i I_i^r + \mu\beta_i I_i^u - \frac{E_i}{Z} \tag{S10}$$

$$\frac{d}{dt}I_i^r = \alpha_i \frac{E_i}{Z} - \frac{I_i^r}{D} \tag{S11}$$

$$\frac{d}{dt}I_i^u = (1-\alpha_i)\frac{E_i}{Z} - \frac{I_i^u}{D} \tag{S12}$$

Consider $X = [E_i, I_i^r, I_i^u]^T$ as infected states. Let $f_i$ be the rate of appearance of new infections in state $i$ and $v_i$ be other transitions among the states such that:

$$f = \begin{pmatrix} \beta_i I_i^r + \mu\beta_i I_i^u \\ 0 \\ 0 \end{pmatrix} \quad \text{and} \quad v = \begin{pmatrix} \frac{E_i}{Z} \\ \frac{I_i^r}{D} - \alpha_i \frac{E_i}{Z} \\ \frac{I_i^u}{D} - (1-\alpha_i)\frac{E_i}{Z} \end{pmatrix}.$$

Given the disease-free equilibrium $\mathcal{E}_i = [N_i, 0, 0, 0, 0]^T$, we have

$$\mathbf{F} = \frac{\partial f}{dX}\Big|_{\mathcal{E}_i} = \begin{pmatrix} 0 & \beta_i & \mu\beta_i \\ 0 & 0 & 0 \\ 0 & 0 & 0 \end{pmatrix} \quad \text{and} \quad \mathbf{V} = \frac{\partial v}{dX}\Big|_{\mathcal{E}_i} = \begin{pmatrix} \frac{1}{Z} & 0 & 0 \\ -\frac{\alpha_i}{Z} & \frac{1}{D} & 0 \\ -\frac{1-\alpha_i}{Z} & 0 & \frac{1}{D} \end{pmatrix}$$

where $\mathbf{F}$ is nonnegative and $\mathbf{V}$ is nonsingular. Let the NGM $\mathbf{G} = \mathbf{F}\mathbf{V}^{-1}$, hence $Rt_i$ is the dominant eigenvalue of $\mathbf{G}$, i.e.,

$$Rt_i = \alpha_i\beta_i D + (1-\alpha_i)\mu\beta_i D$$

#### S1.4 Piecewise SEIR model

Table S2: Split data according to wave variants for piecewise SEIR model

| **Variant wave** | **Time period** | **Country** |
|---|---|---|
| Alpha | 2/12/2020 to 25/03/2021 | Mozambique |
| Delta | 2/06/2021 to 10/10/2021 | Mozambique |
| Omicron BA.1 | 1/11/2021 to 28/03/2022 | Mozambique |
| Omicron BA.1 | 17/11/2021 to 26/01/2022 | Zimbabwe |

#### S1.5 State variables and parameter updates

$$O^i_{t,post} = \frac{\sigma^2_{t,obs}}{\sigma^2_{t,obs} + \sigma^2_{t,prior}}\bar{O}_{t,prior} + \frac{\sigma^2_{t,prior}}{\sigma^2_{t,obs} + \sigma^2_{t,prior}}O_t + \sqrt{\frac{\sigma^2_{t,obs}}{\sigma^2_{t,obs} + \sigma^2_{t,prior}}}(O^i_{t,prior} - \bar{O}_{t,prior}), \tag{S13}$$

where, the posterior and prior of the observed variable for the $i^{th}$ ensemble member at time $t$ is $O^i_{t,post}$ and $O^i_{t,prior}$, respectively; $\bar{O}_{t,prior}$ is the mean of the prior observed variable; $\sigma^2_{t,obs}$ and $\sigma^2_{t,prior}$ are the variances of the observation and the prior observed variable, respectively; and $O_t$ is the observation at time $t$.

$$x^i_{t,post} = x^i_{t,prior} + \frac{\sigma\big(\{x_{t,prior}\}_n, \{O_{t,prior}\}_n\big)}{\sigma^2_{t,prior}}\big(O^i_{t,post} - O^i_{t,prior}\big), \tag{S14}$$

where, $x^i_{t,post}$ and $x^i_{t,prior}$ are the posterior and prior of the unobserved variable or parameter for the $i^{th}$ ensemble member at time $t$; $\sigma\big(\{x_{t,prior}\}_n, \{O_{t,prior}\}_n\big)$ is the covariance between the prior of the unobserved variable/parameter $\{x_{t,prior}\}_n$ and the prior of the observed variable $\{O_{t,prior}\}_n$ at time $t$.

#### S1.6 Initial priors

Table S3: Prior ranges (all assumed uniformly distributed) from which initial conditions for each ensemble member were randomly selected

| | Prior range | Source/ Rationale |
|---|---|---|
| **Variables** | | |
| $S_i$ | $[0.9N_i,\ N_i]$ (Alpha)<br>$[0.75N_i,\ 0.95N_i]$ (Delta)<br>$[0.3N_i,\ 0.85N_i]$ (Omicron) | a large proportion of the population in province $i$ is susceptible initially. See Section 2.2.2 for detailed explanation. |
| $E_i$ | $[0,\ 0.0001N_i]$ | [8] |
| $I^r_i$ | [0, 0] | |
| $I^u_i$ | [0, 0] | |
| **Parameters** | | |
| $\beta_i$ | [0.5, 2.0] | based on previous estimates from [6]. |
| $\mu$ | 0.8 | based on estimates from [6] [5]. |
| $Z$ | 4 | based on previous estimates from [6] |
| $\alpha_i$ | [0.004 (varied), 0.1] | based on a study by Han *et al.* [4] and Evans *et al.* [3]. The lower bound was varied for the provinces (see Section 2.2.2 for detailed explanation) |
| $D$ | 4 | based on previous estimates from [6] |

#### S1.7 Filter divergence

$$\mathbf{x}_t = \bar{\mathbf{x}}_{t-1} + \lambda(\bar{\mathbf{x}} - \bar{\mathbf{x}}_{t-1}), \tag{S15}$$

where $\mathbf{x}_t$ is the prior and $\bar{\mathbf{x}}_{t-1}$ is the mean over ensemble members $\mathbf{x}_{t-1}$.

**Algorithm S1** EAKF for estimation of model parameters

set OEV using Equation (2.2)
**Initialize:** at $t = 0$ generate 300-member ensemble using LHS technique from prior ranges
run the stochastic SEIR metapopulation model, Equation (S1)–Equation (S4) for 1 day to initialize the system
**for** $t = 1$ to $T$ **do**
  obtain posterior from time $t - 1$
  inflation of state variables and estimated parameters using Equation (S15)
  run the model and obtain the priors
  **for** $i = 1$ to $L$(number of locations) **do**
    update observable state variables using Equation (S13)
    update unobservable local state variables and global parameters using Equation (S14). Update local parameters using the Kalman gain for each province
    re-initialize individual ensemble members if parameter posteriors migrate outside prior bounds or state variables $< 0$
  **end for**
**end for**

#### S1.8 Gravity model test to estimate human mobility

Due to the absence of inter-provincial human mobility data for both Mozambique and Zimbabwe, we explored use of a gravity model to estimate human movement. Gravity models have been applied to other infectious diseases such as cholera in Haiti [9], Ebola virus disease in Sierra Leonne [12] and prevaccination epidemics of measles in England and Wales [11]. A gravity model assumes that movement between two places is directly proportional to population size and inversely proportional to distance. However, multiple experiments with gravity model formulations produced poor representation of COVID-19 in Mozambique and Zimbabwe, likely due to a poor match to baseline movement dynamics and the effects of lock-downs implemented in both countries during the COVID-19 outbreak. Hence, we implemented the metapopulation model without movement between provinces.

### S2 Results

#### S2.1 Data sources

##### COVID-19 data

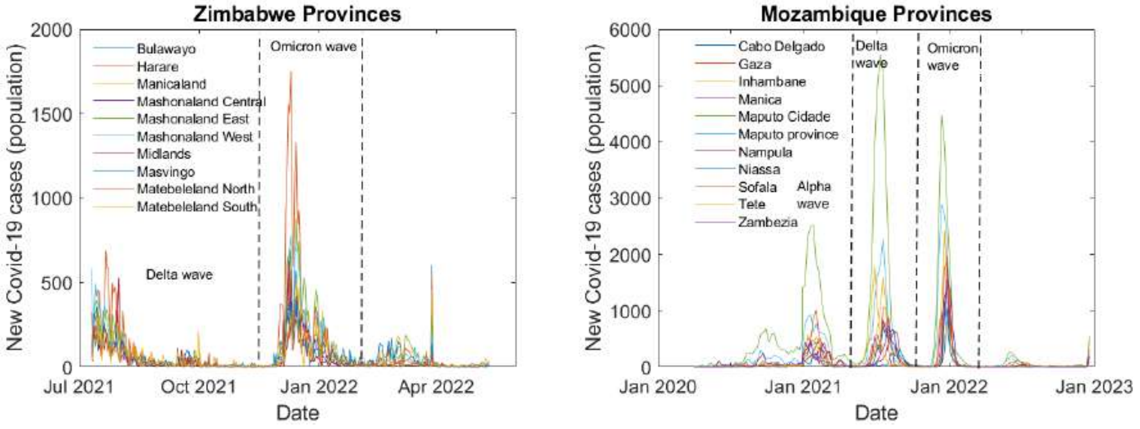


(a) Zimbabwe daily reported cases. (b) Mozambique weekly reported cases

Figure S1: Time series of COVID-19 infections in Mozambique and Zimbabwe provinces during the first two years of the pandemic, according to data released by the Mozambique Ministry of Health-SIGILIA and DISA [7] and Zimbabwe COVID-19 hub [1]. The dotted vertical lines indicate the periods for the SARS-CoV-2 variant waves.

Table S4: Cumulative infected reported population across the variant waves for Mozambique (data from [7]).

| **Province (Population)** | **Alpha (total infected) %** | **Delta (total infected) %** | **Omicron BA.1 (total infected) %** |
|---|---|---|---|
| Cabo Delgado (2.3M) | (2978) 0.12 | (4577) 0.2 | (8089) 0.34 |
| Gaza (1.4M) | (3697) 0.26 | (8920) 0.63 | (17290) 1.22 |
| Inhambane (1.5M) | (3948) 0.27 | (10563) 0.71 | (18707) 1.26 |
| Manica (1.9M) | (2233) 0.11 | (6442) 0.33 | (11331) 0.58 |
| Maputo Cidade (1.1M) | (26364) 2.35 | (57662) 5.14 | (78346) 6.99 |
| Maputo (2M) | (10084) 0.51 | (21114) 1.07 | (34177) 1.74 |
| Nampula (5.8M) | (3230) 0.06 | (7054) 0.12 | (11700) 0.2 |
| Niassa (1.8M) | (2363) 0.13 | (7400) 0.41 | (9950) 0.55 |
| Sofala (2.2M) | (5571) 0.24 | (9285) 0.41 | (15845) 0.7 |
| Tete (2.6M) | (2612) 0.1 | (8784) 0.33 | (11244) 0.42 |
| Zambezia (5.1M) | (4569) 0.09 | (8273) 0.16 | (13588) 0.26 |

Table S5: Cumulative infected reported population across the variant wave for Zimbabwe (data from [1]).

| **Province (Population)** | **Omicron BA.1 (total infected) %** |
|---|---|
| Bulawayo (0.7M) | (6904) 1.04 |
| Harare (2.4M) | (18513) 0.76 |
| Manicaland (2M) | (10959) 0.54 |
| Mashonaland Central (1.4M) | (4838) 0.35 |
| Mashonaland East (1.7M) | (14248) 0.82 |
| Mashonaland West (1.9M) | (11986) 0.63 |
| Midlands (1.8M) | (6252) 0.34 |
| Masvingo (1.6M) | (7195) 0.44 |
| Matebeleland North (0.8M) | (7560) 0.91 |
| Matebeleland South (0.8M) | (5841) 0.77 |

#### S2.2 Validation of the model-inference system using sythetic outbreaks

Table S6: Combinations of model parameter values used to generate synthetic outbreaks

| **Outbreak#** | $\beta_\mathbf{i}[\mathbf{0.8}, \mathbf{2.0}]$ | $\alpha_\mathbf{i}[\mathbf{0.004}, \mathbf{0.1}]$ |
|---|---|---|
| $1-3$ | 0.8 | 0.01 & 0.04 & 0.09 |
| $4-6$ | 1.3 | 0.01 & 0.04 & 0.09 |
| $7-9$ | 1.8 | 0.01 & 0.04 & 0.09 |

#### S2.3 Synthetic testing accuracy of parameter estimation

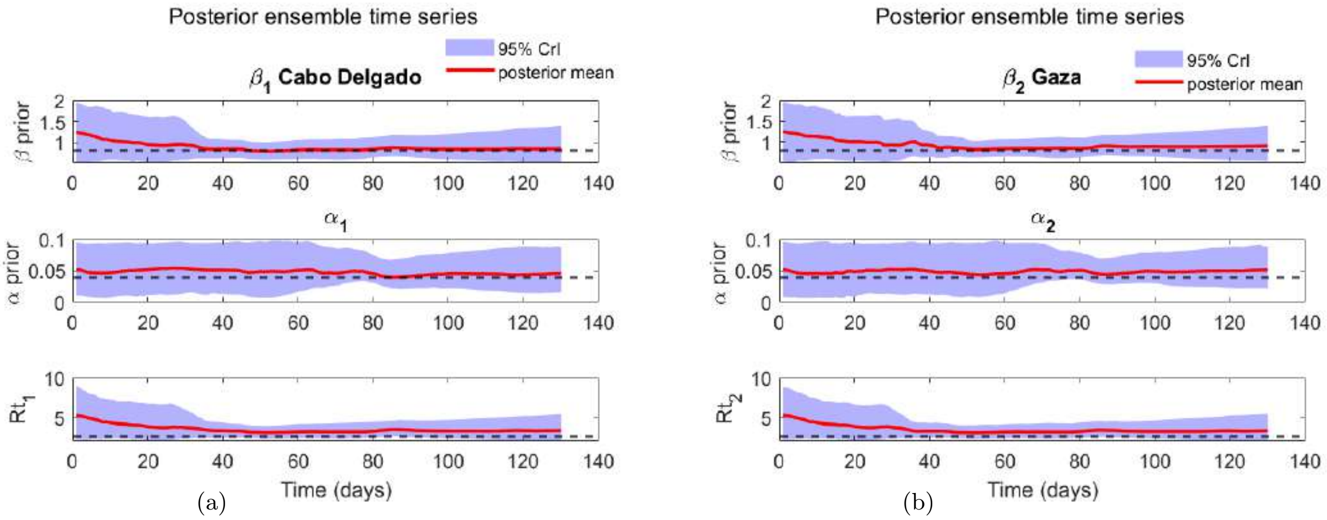


(a) (b)

Figure S2: **Evolution of parameter posteriors over time.** The dashed black lines represent the truth values, the uncertainties around the mean values (red line) are filled in blue, and the y-axis represent the initial prior parameter ranges (described in Table S3).

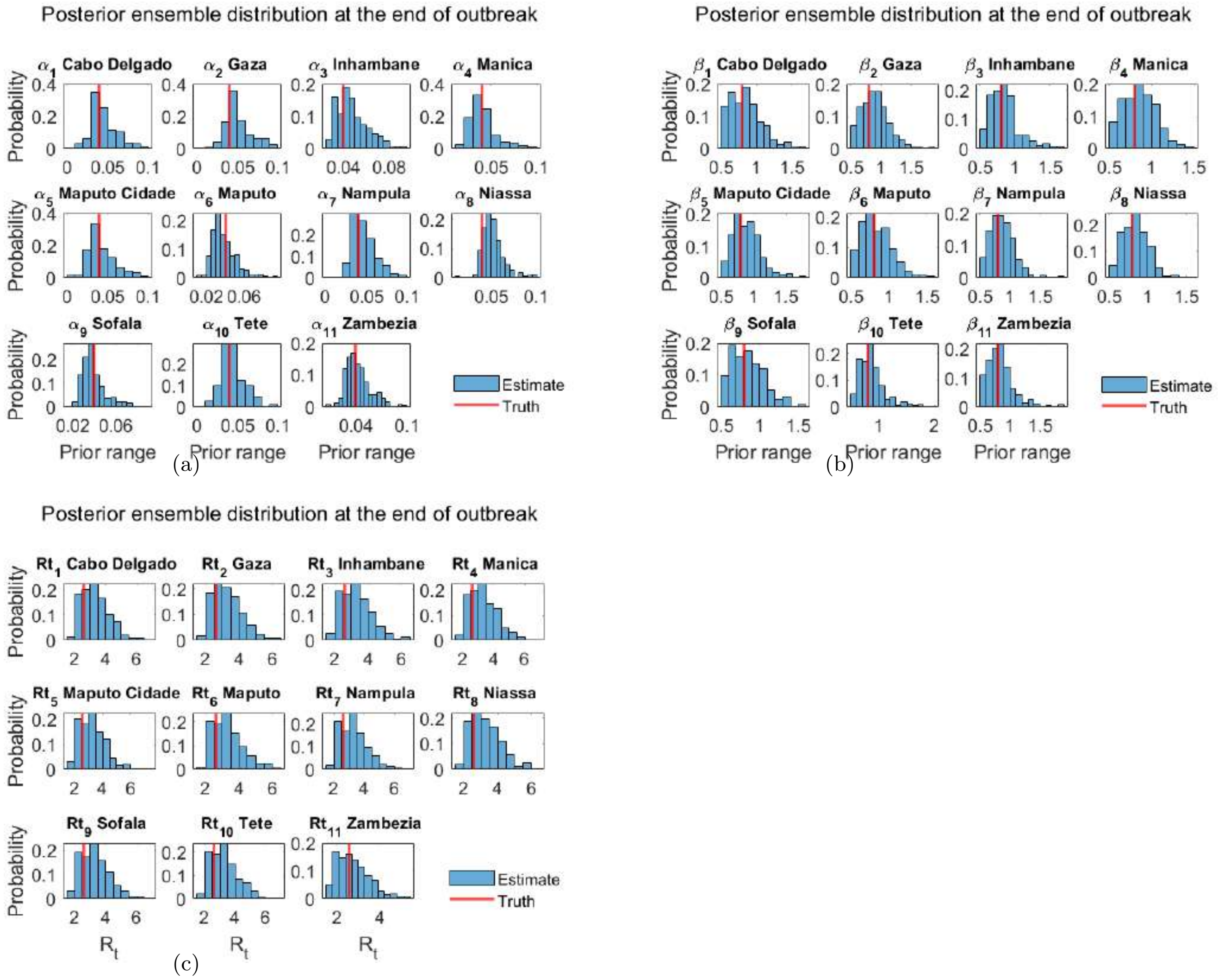


Figure S3: **Parameter estimation accuracy** using truth values, truth $\beta_i$ =0.8, truth $\alpha_i$ =0.04, $\mu$ = 0.8, D=Z=4. The vertical red lines represent the truth values, the blue bars represent the distribution of the posterior parameter estimates, and the x-axis depicts the initial prior parameter ranges (described in Table S3).

#### S2.4 Time series posterior parameter estimates

##### S2.4.1 Mozambique provinces

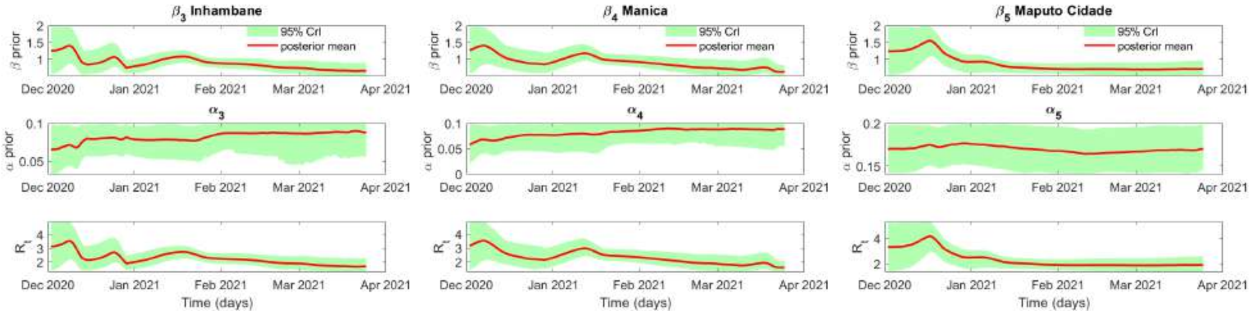


(a) (b) (c)

Figure S4: **Evolution of parameter posteriors over time for Mozambique Alpha wave.** The y-axis represent the initial prior parameter ranges (described in Table S3).

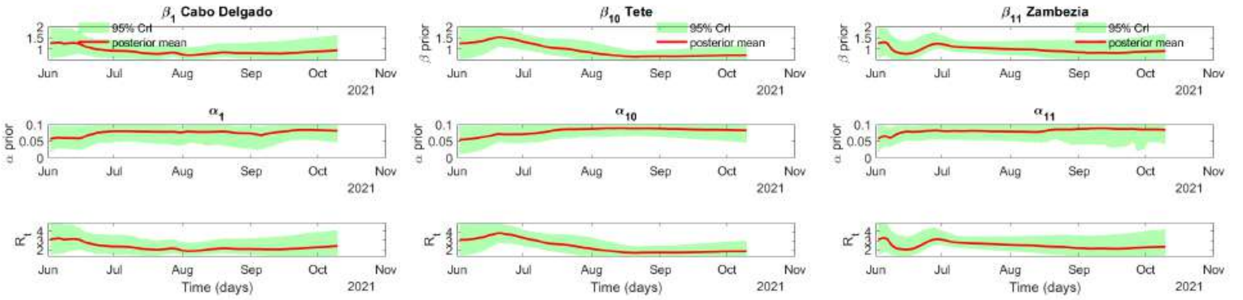


(a) (b) (c)

Figure S5: **Evolution of parameter posteriors over time for Mozambique Delta wave.** The y-axis represent the initial prior parameter ranges (described in Table S3).

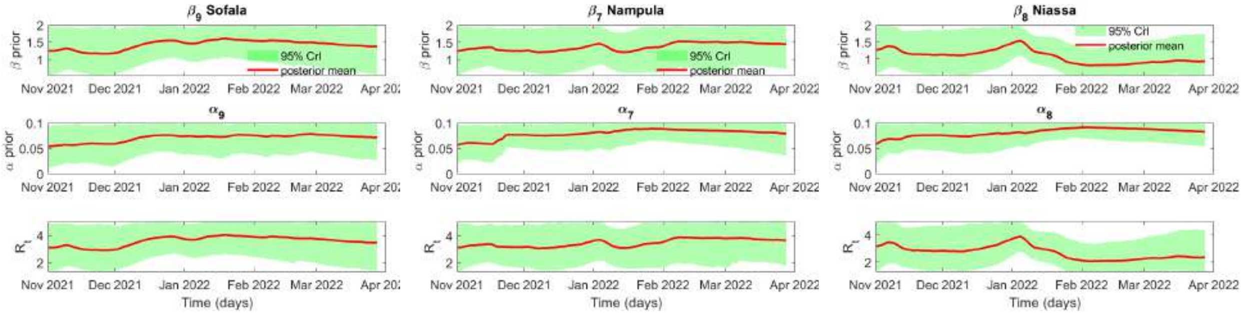


(a) (b) (c)

Figure S6: **Evolution of parameter posteriors over time for Mozambique Omicron (BA.1) wave.** The y-axis represent the initial prior parameter ranges (described in Table S3).

#### S2.5 Posterior fits

##### S2.5.1 Mozambique provinces

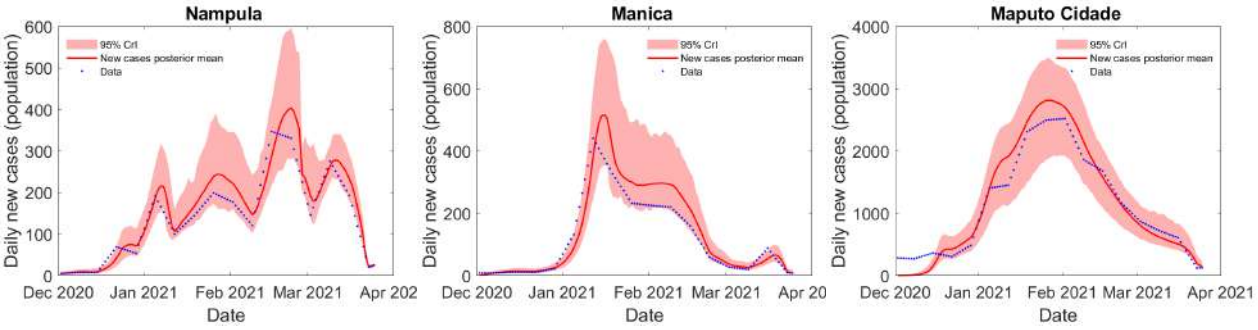


(a) (b) (c)

Figure S7: **Accuracy of curve fitting.** Alpha wave new reported cases from the Mozambique provincial data [7] and model fit.

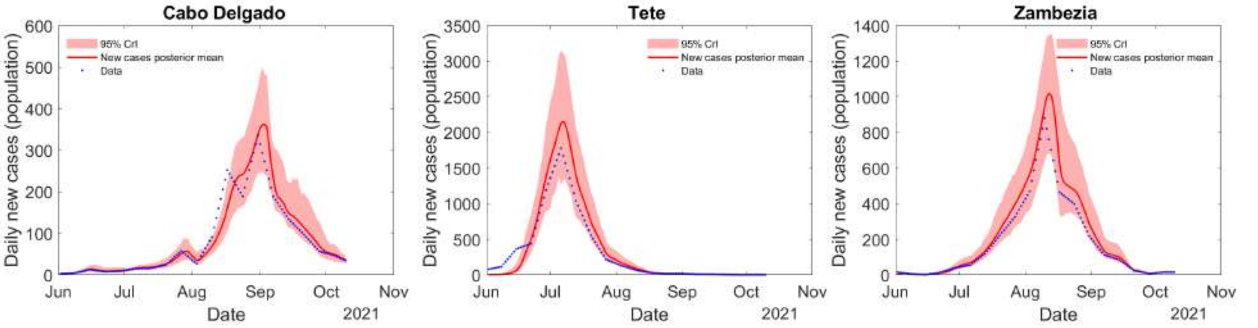


(a) (b) (c)

Figure S8: **Accuracy of curve fitting.** Delta wave new reported cases from the Mozambique provincial data [7] and model fit.

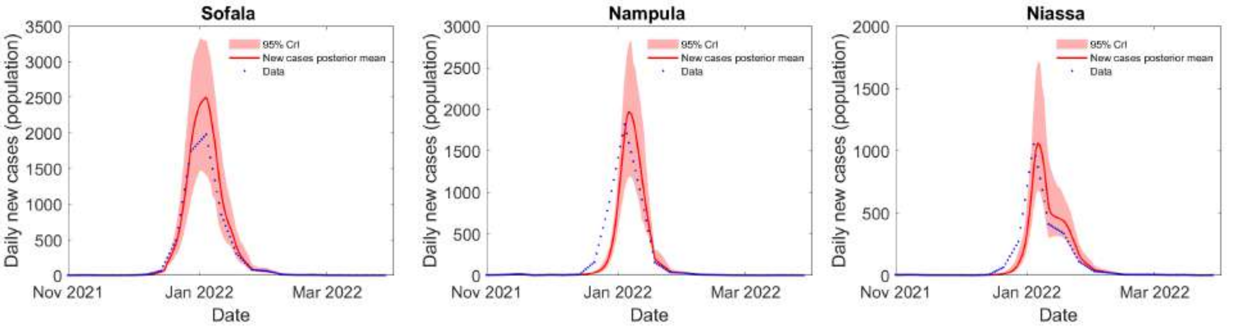


(a) (b) (c)

Figure S9: **Accuracy of curve fitting.** Omicron (BA.1) wave new reported cases from the Mozambique provincial data [7] and model fit.

##### S2.5.2 Zimbabwe provinces

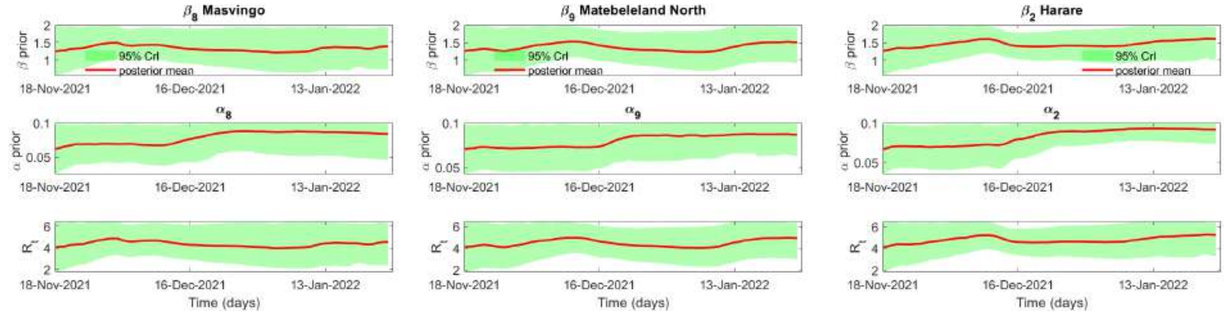


(a) (b) (c)

Figure S10: **Evolution of parameter posteriors over time for Zimbabwe Omicron (BA.1) wave.** The y-axis represent the initial prior parameter ranges (described in Table S3).

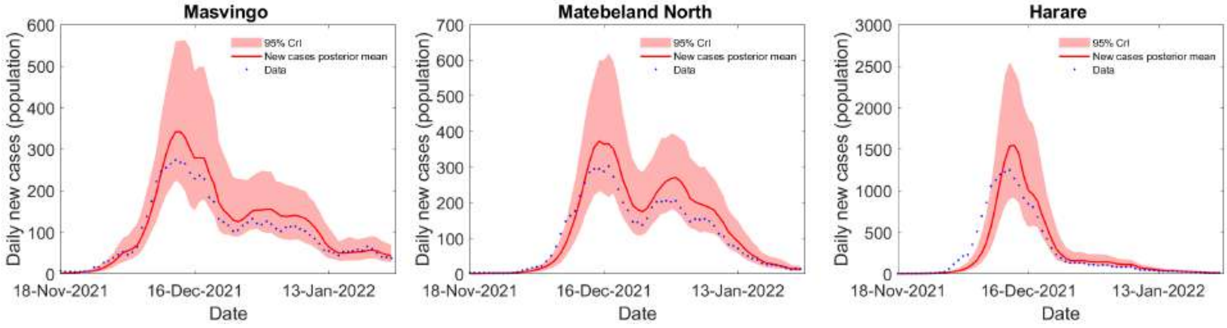


(a) (b) (c)

Figure S11: **Accuracy of curve fitting.** Omicron (BA.1) wave new reported cases from the Zimbabwe provincial data [1] and model fit.

#### S2.6 Distributions of Susceptible depletion mean estimates at the end of each wave

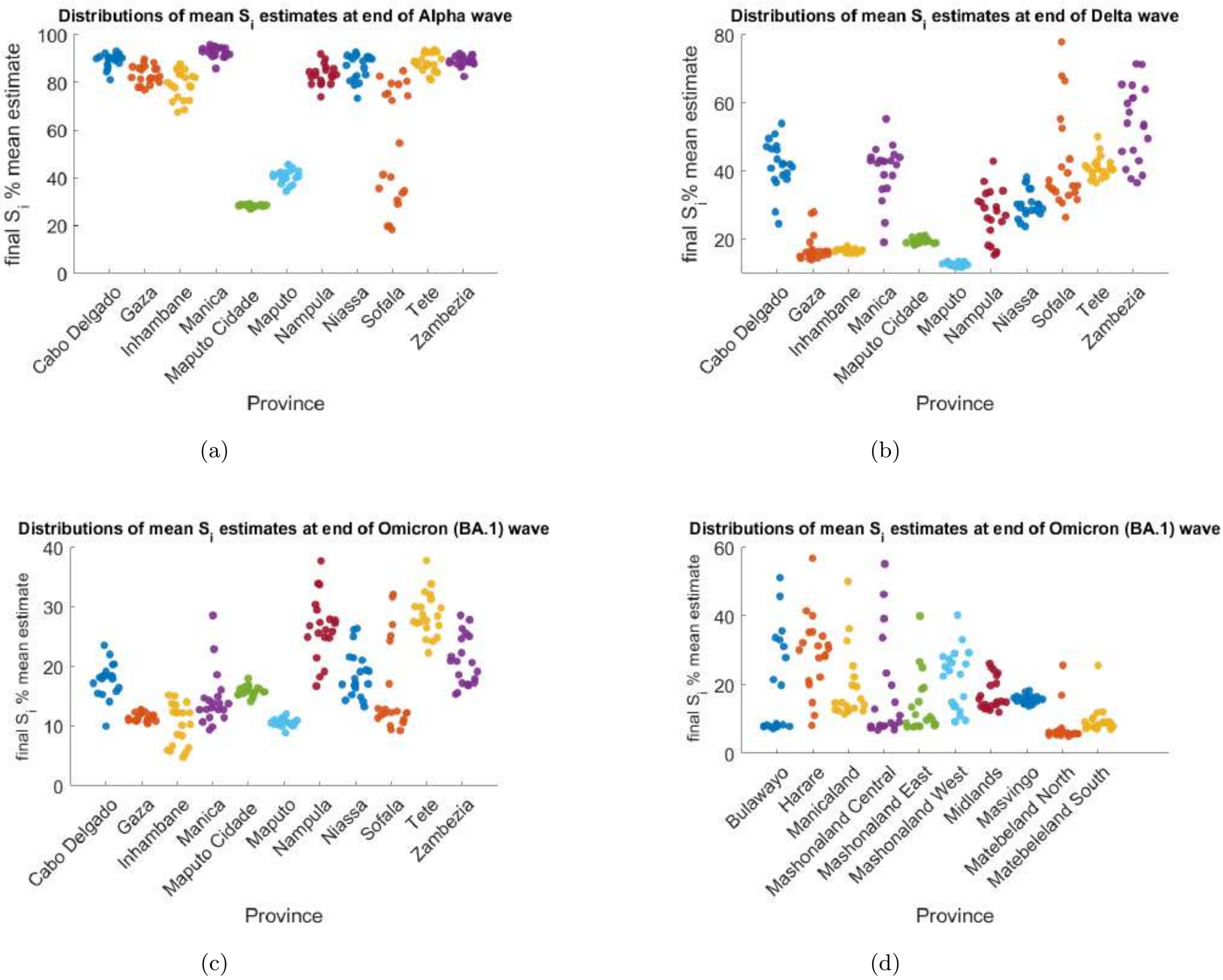


Figure S12: Distribution of the final $S_i$ mean estimates at the end of variant outbreaks (20 runs) for the 11 Mozambique and 10 Zimbabwe provinces.

[7] Ministry of Health-SIGILIA and DISA. Covid-19 cases. Mozambique Ministry of Health.
